# High interleukin-8 serum levels independently predict long-term all-cause mortality in chronic heart failure

**DOI:** 10.1101/2025.08.03.25332911

**Authors:** Jan Traub, Michael K Schuhmann, Ulrich Hofmann, Gustavo Ramos, Niklas Beyersdorf, Stefan Frantz, Stefan Störk, Guido Stoll, Anna Frey

**Affiliations:** Department of Internal Medicine I, University Hospital Würzburg, Würzburg, Germany; German Comprehensive Heart Failure Center, University Hospital Würzburg, Würzburg, Germany; Department of Neurology, University Hospital Würzburg, Würzburg, Germany; Institute for Virology and Immunobiology, University of Würzburg, Würzburg, Germany

**Keywords:** Heart failure, inflammation, cytokines, interleukin-8 (IL-8), mortality, biomarker

## Abstract

**Background:** Chronic heart failure (HF) is a complex syndrome with significant morbidity and mortality, where inflammation is increasingly recognized as a critical factor in its progression. This study investigated the association of cytokine and chemokine profiles with mild cognitive impairment (MCI) and long-term mortality in patients with chronic HF.

**Methods:** Serum concentrations of 13 cytokines were measured in 145 patients from the Cognition.Matters-HF study baseline cohort using a bead-based multiplex assay. Detailed clinical and cognitive evaluations were conducted, and survival data were tracked over 10 years. Cox proportional hazards regression and logistic regression models were applied to assess independent associations with mortality and MCI.

**Results:** No cytokine was independently associated with MCI. However, high interleukin-8 (IL-8) levels (>5.3 pg/ml) were significantly and independently associated with all-cause mortality (HR 2.30; 95% CI 1.30-4.07; p=0.004). After adjusting for age and sex, IL-8 remained a strong predictor (Adj. HR 2.28; 95% CI 1.29-4.05; p=0.005). Further adjustments for clinical and biochemical variables showed that IL-8 (Adj. HR 3.31; 95% CI 1.61-6.79; p=0.001) was the only cytokine independently associated with mortality. In the final multivariable model, IL-8 (Adj. HR 2.57; 95% CI 1.44-4.57; p=0.001) remained a significant predictor along with age, body mass index, left atrial volume index, and six-minute walk distance. Other cytokines, including IL-6 and tumor necrosis factor-α, showed no independent associations with mortality.

**Conclusions:** These findings suggest IL-8’s unique role in HF pathophysiology and its potential as a biomarker and therapeutic target. Further research is needed to validate these results and explore the clinical utility of IL-8 modulation in HF management.

## INTRODUCTION

Heart failure (HF) is a complex clinical syndrome characterized by impaired cardiac function and associated with significant morbidity and mortality. Despite advances in its management, HF remains a leading cause of hospitalization and reduced quality of life, with substantial implications for public health. In recent years, there has been growing interest in understanding the role of inflammation in HF pathophysiology [1]. Myocardial inflammation involves the activation of both innate and adaptive immune systems, leading to tissue injury, maladaptive left ventricular remodeling, and worsening cardiac function [2]. The interplay between systemic cytokines (e.g. interleukin (IL)-1β and tumor necrosis factor (TNF)-α) and chemokines (e.g. monocyte chemoattractant protein-1 (MCP-1)) plays a critical role in recruiting immune cells to the heart, contributing to both acute and chronic phases of inflammation [3]. The complexity of involved inflammatory pathways, difficulty in targeting specific cytokines without triggering unintended side effects, and the diverse etiologies of HF explain the lack of an approved therapy for inflammation in HF.

Given the growing evidence of interleukins’ role in HF and their association with various systemic outcomes, this study aimed to investigate whether specific cytokine profiles are independently associated with mild cognitive impairment (MCI) and long-term mortality in a well-characterized HF cohort.

MCI is a prevalent but often under recognized complication of HF with prognostic implication [4]. Previous reports have associated elevated levels of IL-6 with cognitive decline in HF [5]. However, it remains unclear if this observation is independent of other clinical factors and if other cytokines may also be involved.

Against this backdrop, the observational Cognition.Matters-HF study [6] provides the unique opportunity to explore these relationships as it offers detailed clinical, cognitive, and survival data collected from a well-characterized cohort of HF patients. Thus, in this post-hoc multiplex cytokine analysis, we hypothesized that high cytokine or chemokine levels are independently associated with instant MCI and long-term mortality.

## METHODS

### Cognition.Matters-HF study

This post-hoc analysis based on biomaterial (serum) available from the prospective, investigator-initiated, single-center follow-up Cognition.Matters-HF study, which included HF patients between 2011 and 2014. Approved by the local ethics committee (#245/10) and adhering to the Declaration of Helsinki, all participants provided written informed consent. The study included adult patients with clinically confirmed chronic HF, as defined by the European Society of Cardiology guidelines at the time of enrollment, with a wide range of clinical profiles, including varying levels of risk. Eligible patients were consecutively recruited from the outpatient clinic of the Comprehensive Heart Failure Center Würzburg, with initial screening performed via echocardiography. Exclusion criteria included new-onset or acutely decompensated HF, a history of clinical stroke, psychiatric disorders (such as depression or dementia), carotid artery stenosis greater than 50%, or the presence of implants or devices incompatible with brain MRI.

Baseline and follow-up assessments (at 12, 36, and 60 months) included comprehensive evaluations by a cardiologist, neuropsychologist, neurologist, and neuroradiologist, with efforts to complete all procedures within two days per visit. Clinical examinations, echocardiography, and the 6-minute walk test were conducted according to standard protocols at the Comprehensive Heart Failure Center Würzburg. In august 2024, resident registration offices provided overall 10-year mortality data.

### Blood sampling

Venous blood samples were collected from all patients without the requirement for fasting, as part of routine clinical chemistry assessments conducted at the certified facility of the University Hospital Würzburg [7]. Before the blood draw, participants remained seated for at least 5 minutes. Serum samples were processed promptly, kept at room temperature for 30 minutes, and centrifuged at 2000g for 10 minutes. The serum was then aliquoted into specialized fluid tissue tubes (Micronic, Lelystad, Netherlands) and stored at −80°C in a standardized interdisciplinary biobank until analysis.

### Cytokine measurements

The cytokine measurements were performed using the LEGENDplex™ Human Inflammation Panel 1 (Biolegend®, San Diego, CA), a bead-based multiplex assay designed for simultaneous quantification of 13 human inflammatory cytokines and chemokines. This assay uses fluorescence-encoded beads, each conjugated with specific antibodies targeting the cytokines of interest. Serum samples were mixed with the antibody-coated beads and incubated to allow binding. Subsequently, biotinylated detection antibodies specific to the bound cytokines were added, followed by Streptavidin-Phycoerythrin (SA-PE), which binds to the detection antibodies. The fluorescent signals emitted by the SA-PE were proportional to the cytokine concentrations and were quantified using a flow cytometer. A standard curve generated within the same assay was used for determining the cytokine levels in the samples.

### Cognitive testing

Extensive neuropsychological testing included the Test Battery of Attentional Performance [8] for the domains of intensity and selectivity of attention. The Visual and Verbal Memory Test [9] was used to assess visual/verbal memory performance, while working memory was assessed using the Digit Span Forward and Block Tapping Span Forward tests [10]. The Regensburg Word Fluency Test [11] and the HAMASCH 5-point test [12] were used to assess visual/verbal fluency. These tests were validated in healthy volunteers to account for the modifying effect of age, sex and level of education. These factors were taken into account in the T-standardized performance scores with a mean of 50 and a standard deviation of 10 as the mean reference comparator. MCI was defined as underperformance (T score <40) in at least one of the assessed domains.

### Statistical analysis

Data were reported as medians with interquartile ranges or as counts with percentages, depending on the variable type. Statistical analyses included T-tests for comparing means of normally distributed variables and Mann-Whitney U tests for those not following a normal distribution. Categorical data were analyzed using Chi-squared tests or Fisher’s exact tests as appropriate. Cox regression analyses were conducted to evaluate the impact of various predictors on survival outcomes, providing hazard ratios and p-values. Kaplan-Meier plots were created to illustrate survival distributions, and log-rank tests were applied to assess differences between groups.

Two multivariable models were used to assess the association of cytokines with the outcomes. Model 1 included adjustments for age and sex as baseline demographic variables. Model 2 incorporated additional clinical and biochemical variables to account for potential confounders, including body mass index, C-reactive protein, leukocyte count, estimated glomerular filtration rate, hemoglobin A1c, hemoglobin, urea, uric acid, low-density lipoprotein, alanine aminotransferase, aspartate aminotransferase, albumin, atrial fibrillation, arterial hypertension, diabetes mellitus, hyperlipidemia, smoking history, coronary artery disease, history of myocardial infarction, creatine kinase, NT-proBNP, left ventricular ejection fraction, left atrial volume index, New York Heart Association functional class, six-minute walk distance, left ventricular end-diastolic volume, E/e’ ratio, deceleration time, and isovolumetric relaxation time. The incremental inclusion of these variables in Model 2 allowed for more comprehensive adjustment and evaluation of independent associations.

## RESULTS

### Patient characteristics

Sufficient biomaterial (350µl serum) for this post-hoc cytokine quantification for was available from 145 patients (98%) within the Cognition.Matters-HF baseline cohort. As shown in **Table 1**, their median age was 64 years (interquartile range 56-72 years) and 22 patients (15%) were female. The majority of HF patients suffered from ischemic HF (66%), was in Ney York Heart Association (NYHA) functional class II (60%) and had HF with midrange ejection fraction (50%). Accordingly, median left ventricular ejection fraction (LVEF) was 44 % (38-48) and NT-proBNP was 680 (238-1765). Arterial hypertension was the most common cardiovascular risk factor (79%), followed by hyperlipidemia (72%) and smoking (61%). Due to patient recruitment between the years 2011 and 2014, none of the patients received angiotensin receptor-neprilysin or sodium glucose linked transporter 2 inhibitors. MCI was detected in 101 patients (70%).

**Table 1.**
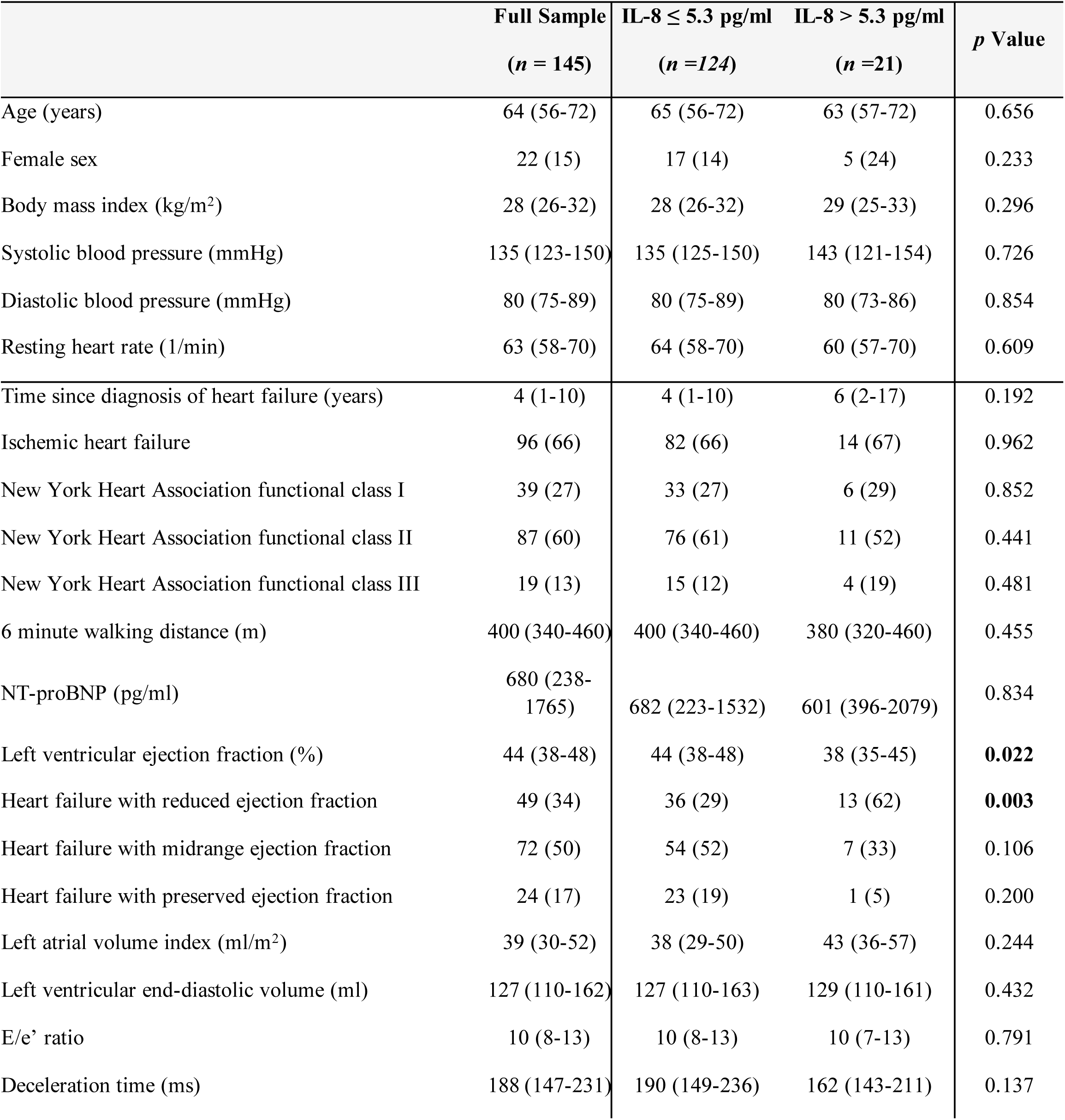
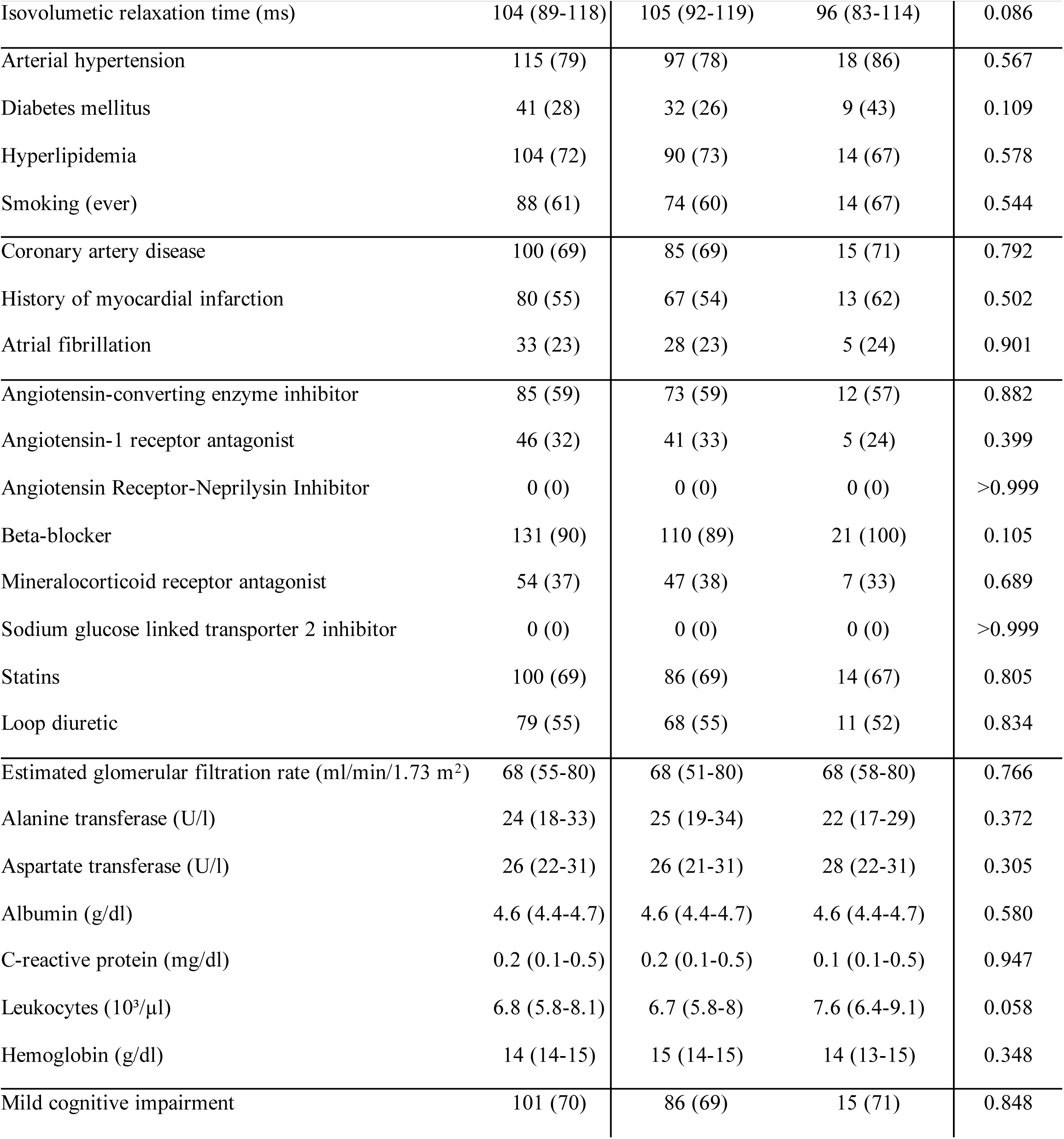
Baseline Characteristics of Heart Failure Patients by IL-8 Levels. Summary of demographic, clinical, and laboratory characteristics of heart failure patients stratified by IL-8 levels (≤5.3 pg/ml vs. >5.3 pg/ml). Data are presented as medians (interquartile range) or counts (%), with p-values for comparisons.

### Cytokine measurements

Absolute values of detected serum concentrations of the 13 quantified cytokines are shown in **Table 2**, including detection limit, median, interquartile range and maximum. Only for Monocyte chemoattractant protein-1 (MCP-1), IL-18 and IL-33, all patients hat concentrations above the detection limit. High cytokine levels (↑) were defined as above detection limit or (if higher) median cytokine concentration. Cytokine concentrations were strongly interrelated with one another, as displayed in Figure 1.

**Figure 1:**
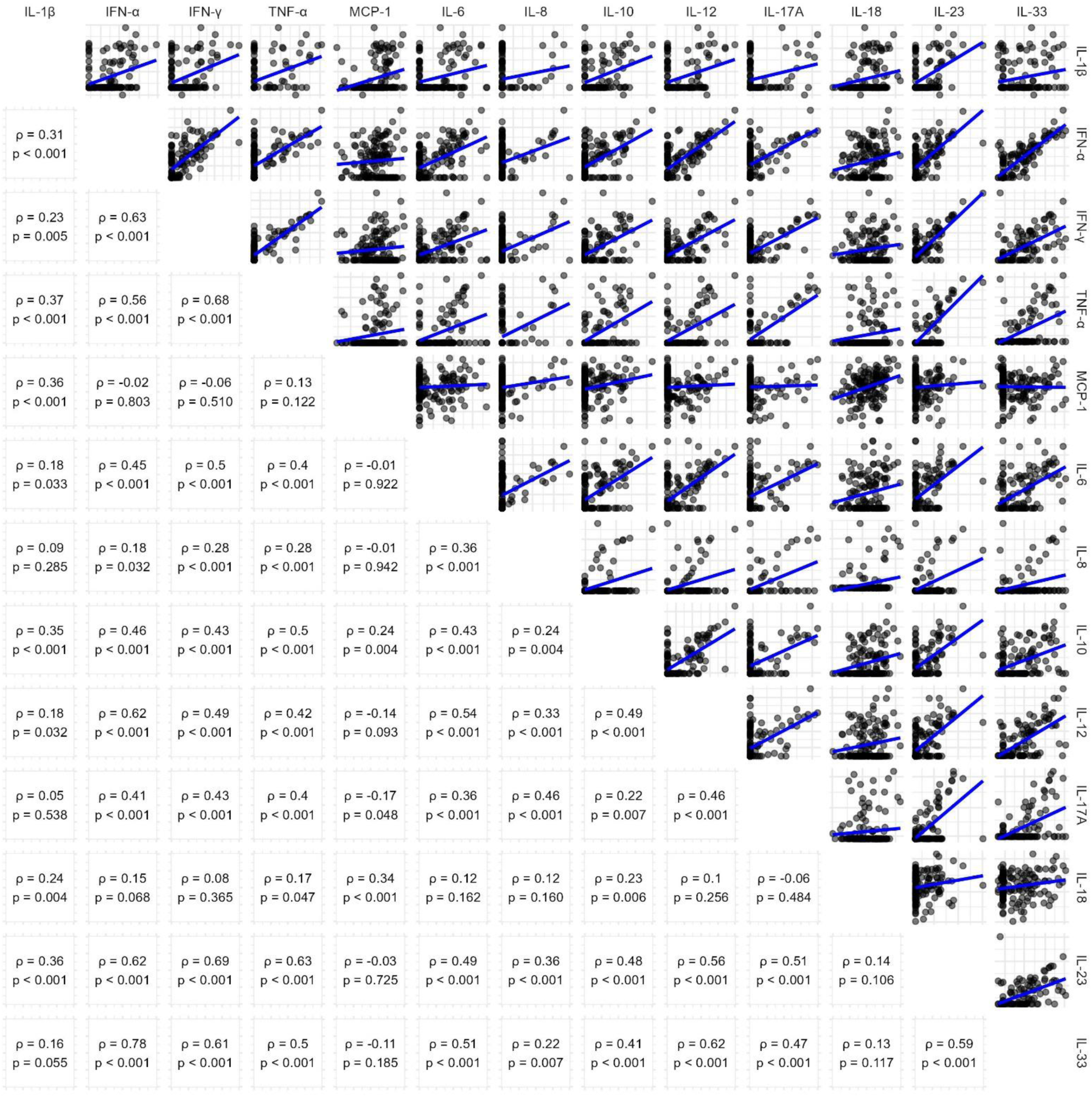
Pairwise Correlation of Cytokine Concentrations. Scatterplots (upper triangle) and Spearman’s correlation coefficients (lower triangle) showing relationships between cytokine levels in the cohort. Log-log scales and linear trendlines illustrate the trends in cytokine interactions.

**Table 2.** Cytokine Levels in the Study Cohort. Median, interquartile range (IQR), and detection limits of 13 cytokines measured in the cohort. High cytokine levels are defined as concentrations above the detection limit or, if higher, the median value.

| Cytokine | Detection limit (DL) | n (%) below DL | n (%) high levels* | median (IQR) | maximum |
| --- | --- | --- | --- | --- | --- |
| Interleukin-1β (pg/ml) | 2.06 | 96 (66) | 49 (34) | 2.1 (2.1-11.5) | 772.5 |
| Interferon-α (pg/ml) | 0.49 | 45 (31) | 74 (50) | 1.2 (0.5-2.7) | 27.8 |
| Interferon-γ (pg/ml) | 1.39 | 59 (41) | 74 (50) | 1.9 (1.4-5.4) | 171.5 |
| Tumor necrosis factor-α (pg/ml) | 2.11 | 109 (75) | 36 (25) | 2.1 (2.1-2.4) | 109.4 |
| Monocyte chemoattractant protein-1 (pg/ml) | 0.24 | 0 (0) | 74 (50) | 334 (205-502) | 1224.9 |
| Interleukin-6 (pg/ml) | 1.35 | 64 (44) | 74 (50) | 1.8 (1.3-7.6) | 68.9 |
| Interleukin-8 (pg/ml) | 5.32 | 124 (86) | 21 (14) | 5.3 (5.3-5.3) | 247.3 |
| Interleukin-10 (pg/ml) | 0.95 | 77 (53) | 68 (47) | 1.0 (1.0-3.9) | 150.6 |
| Interleukin-12 (pg/ml) | 0.56 | 72 (50) | 74 (50) | 0.6 (0.6-2.6) | 54.3 |
| Interleukin-17A (pg/ml) | 0.09 | 96 (66) | 49 (34) | 0.1 (0.1-0.1) | 1.2 |
| Interleukin-18 (pg/ml) | 0.76 | 0 (0) | 74 (50) | 377 (271-511) | 1191.4 |
| Interleukin-23 (pg/ml) | 1.73 | 79 (55) | 66 (46) | 1.7 (1.7-6.5) | 1050.0 |
| Interleukin-33 (pg/ml) | 8.94 | 0 (0) | 74 (50) | 15 (9-41) | 322.6 |

### Logistic regression for mild cognitive impairment

A total of 101 patients suffered from MCI (70%). Univariable logistic regression analysis revealed that none of the analyzed cytokines associated with instant MCI (**Table 3**). This was also the case in multivariable models, which adjusted for age, sex and multiple clinical parameters.

**Table 3.**
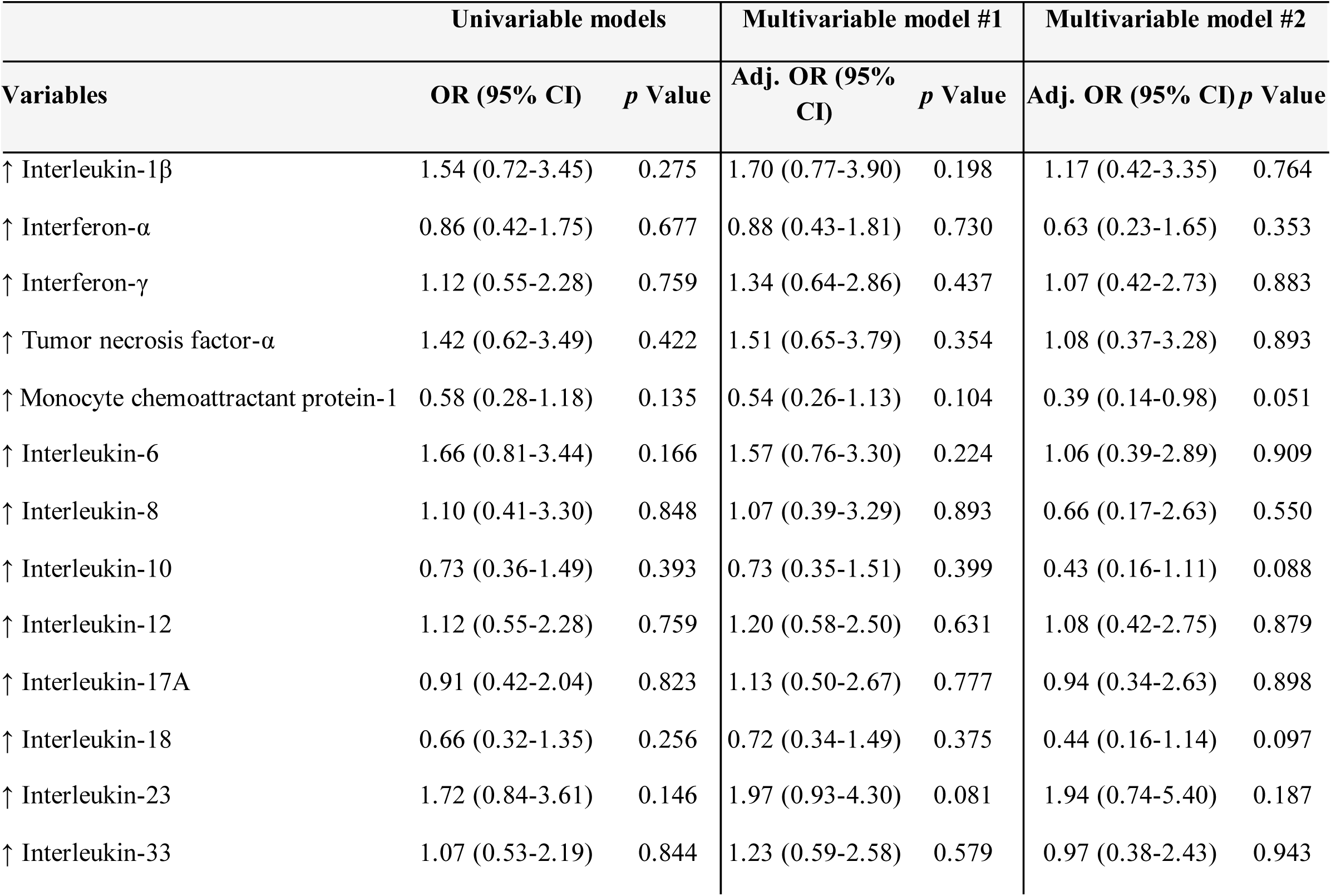
Logistic Regression Models for Mild Cognitive Impairment. Univariable and multivariable logistic regression models showing odds ratios (OR) and 95% confidence intervals (CI) for the association of cytokines with mild cognitive impairment in heart failure patients.

| Univariable models |  |  | Multivariable model #1 |  | Multivariable model #2 |  |
| --- | --- | --- | --- | --- | --- | --- |
| Variables | OR (95% CI) | *p* Value | Adj. OR (95% CI) | *p* Value | Adj. OR (95% CI) | *p* Value |
| ↑ Interleukin-1β | 1.54 (0.72-3.45) | 0.275 | 1.70 (0.77-3.90) | 0.198 | 1.17 (0.42-3.35) | 0.764 |
| ↑ Interferon-α | 0.86 (0.42-1.75) | 0.677 | 0.88 (0.43-1.81) | 0.730 | 0.63 (0.23-1.65) | 0.353 |
| ↑ Interferon-γ | 1.12 (0.55-2.28) | 0.759 | 1.34 (0.64-2.86) | 0.437 | 1.07 (0.42-2.73) | 0.883 |
| ↑ Tumor necrosis factor-α | 1.42 (0.62-3.49) | 0.422 | 1.51 (0.65-3.79) | 0.354 | 1.08 (0.37-3.28) | 0.893 |
| ↑ Monocyte chemoattractant protein-1 | 0.58 (0.28-1.18) | 0.135 | 0.54 (0.26-1.13) | 0.104 | 0.39 (0.14-0.98) | 0.051 |
| ↑ Interleukin-6 | 1.66 (0.81-3.44) | 0.166 | 1.57 (0.76-3.30) | 0.224 | 1.06 (0.39-2.89) | 0.909 |
| ↑ Interleukin-8 | 1.10 (0.41-3.30) | 0.848 | 1.07 (0.39-3.29) | 0.893 | 0.66 (0.17-2.63) | 0.550 |
| ↑ Interleukin-10 | 0.73 (0.36-1.49) | 0.393 | 0.73 (0.35-1.51) | 0.399 | 0.43 (0.16-1.11) | 0.088 |
| ↑ Interleukin-12 | 1.12 (0.55-2.28) | 0.759 | 1.20 (0.58-2.50) | 0.631 | 1.08 (0.42-2.75) | 0.879 |
| ↑ Interleukin-17A | 0.91 (0.42-2.04) | 0.823 | 1.13 (0.50-2.67) | 0.777 | 0.94 (0.34-2.63) | 0.898 |
| ↑ Interleukin-18 | 0.66 (0.32-1.35) | 0.256 | 0.72 (0.34-1.49) | 0.375 | 0.44 (0.16-1.14) | 0.097 |
| ↑ Interleukin-23 | 1.72 (0.84-3.61) | 0.146 | 1.97 (0.93-4.30) | 0.081 | 1.94 (0.74-5.40) | 0.187 |
| ↑ Interleukin-33 | 1.07 (0.53-2.19) | 0.844 | 1.23 (0.59-2.58) | 0.579 | 0.97 (0.38-2.43) | 0.943 |

### Cox regression analysis for overall death

Within 10 years after study inclusion, 62 patients (43%) died of any cause. Using univariable cox proportional hazard models (**Table 4**), significant associations of ↑ IL-6 (HR 2.07; 95% CI 1.30-3.32; p=0.002) and ↑ IL-8 (HR 2.30; 95% CI 1.30-4.07) were identified. Elevations of other cytokines did not associated with overall mortality in this univariable setting (p>0.05).

**Table 4.**
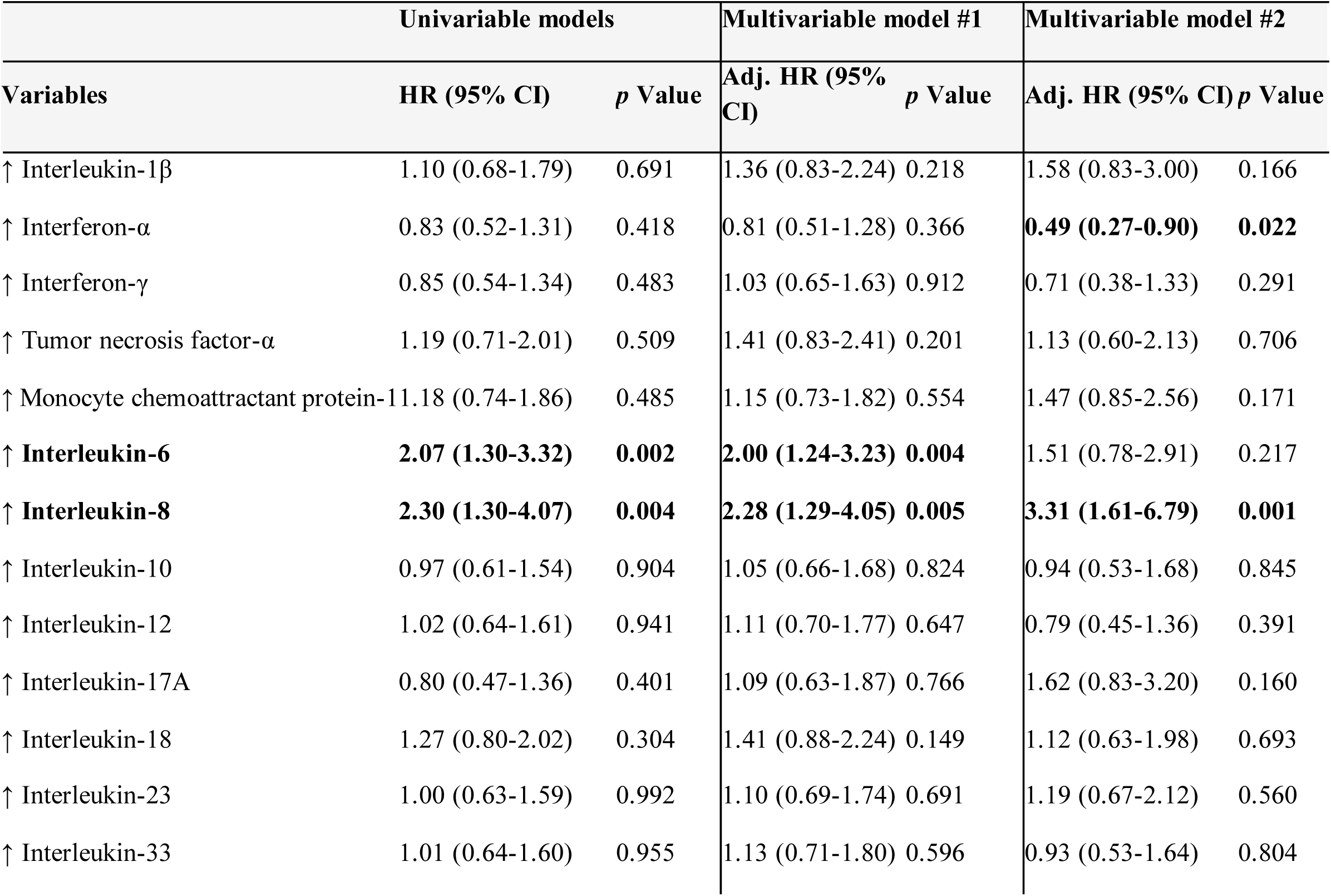
Cox Regression Models for 10-Year Mortality. Univariable and multivariable Cox regression models with hazard ratios (HR) and 95% confidence intervals (CI) for the association of cytokines with all-cause mortality over 10 years in heart failure patients.

In a next step, age and sex were added to perform multivariable cox regression. Like before, only ↑ IL-6 (Adj. HR 2.00; 95% CI 1.24-3.23; p=0.004) and ↑ IL-8 (Adj. HR 2.28; 95% CI 1.29-4.05; p=0.005) significantly predicted all-cause mortality.

To account for other possible confounding factors, additional adjustment for body mass index, C-reactive protein, leukocyte count, estimated glomerular filtration rate, hemoglobin A1c, hemoglobin, urea, uric acid, low-density lipoprotein, alanine aminotransferase, aspartate aminotransferase, albumin, atrial fibrillation, arterial hypertension, diabetes mellitus, hypolipoproteinemia, smoking, coronary artery disease, former myocardial infarction, creatine kinase, NT-proBNP, LVEF, left atrial volume index (LAVI), NYHA class, 6-minute walking distance (6-MWD), left ventricular end diastolic volume, E/e’ ratio, deceleration time and isovolumetic relaxation time was done. Here, ↑ IL-8 (Adj. HR 3.31, 95% CI 1.61-6.79) remained independently associated with mortality, along with ↑ IFN-α (Adj. HR 0.49, 95% CI 0.27-0.90; p=0.022).

To create a final model for overall death (**Table 5**), we performed backward elimination according to p value with a threshold of 0.05 from an initial model, which included ↑ IFN-α and ↑ IL-8, along with all of the above-mentioned possible confounders. Along with age, body mass index, LAVI and 6-MWD, ↑ IL-8 remained an independent predictor of all-cause mortality (Adj. HR 2.54; 95% CI 1.44-4.57); p=0.001). This final model demonstrated good predictive accuracy with a Harrell’s Concordance Index of 0.757, and the likelihood ratio test (χ² = 64, df = 5, p < 0.001), indicating a highly significant relationship between the predictors and survival outcomes.

**Table 5.** Final Multivariable Cox Regression Model for 10-Year Mortality. Results of the final Cox regression model, generated by backward elimination with a p-value threshold of 0.05, including IL-8 levels, clinical, and demographic variables as predictors of all-cause mortality.

| Variables | Adj. HR 95% CI | p Value |
| --- | --- | --- |
| Age (years) | 1.03 (1.00-1.06) | 0.022 |
| Body mass index (kg/m<sup>2</sup>) | 0.93 (0.88-0.99) | 0.014 |
| Left atrial volume index (ml/m<sup>2</sup>) | 1.03 (1.01-1.04) | 0.001 |
| Six-minute walk distance (m) | 0.99 (0.99-1.00) | <0.001 |
| ↑ Interleukin-8 | 2.57 (1.44-4.57) | 0.001 |

*Harrell's Concordance Index: 0.757, Likelihood ratio test $\chi^2(df = 5) = 64, p < 0.001$*

As visualized in **Figure 2**, patients with ↑ IL-8 levels (>5.3 pg/ml) indeed had lower 10-year survival rates (33%) compared to those with IL-8 levels ≤5.3 pg/ml (61%). A log-rank test confirmed a statistically significant difference between both groups (p=0.003). The adjusted survival curve derived from the final Cox proportional hazards model adjusted for relevant covariates. Patients with IL-8 levels above 5.3 pg/ml exhibited significantly worse survival compared to those with lower levels, after controlling for other predictors in the model.

**Figure 2:**
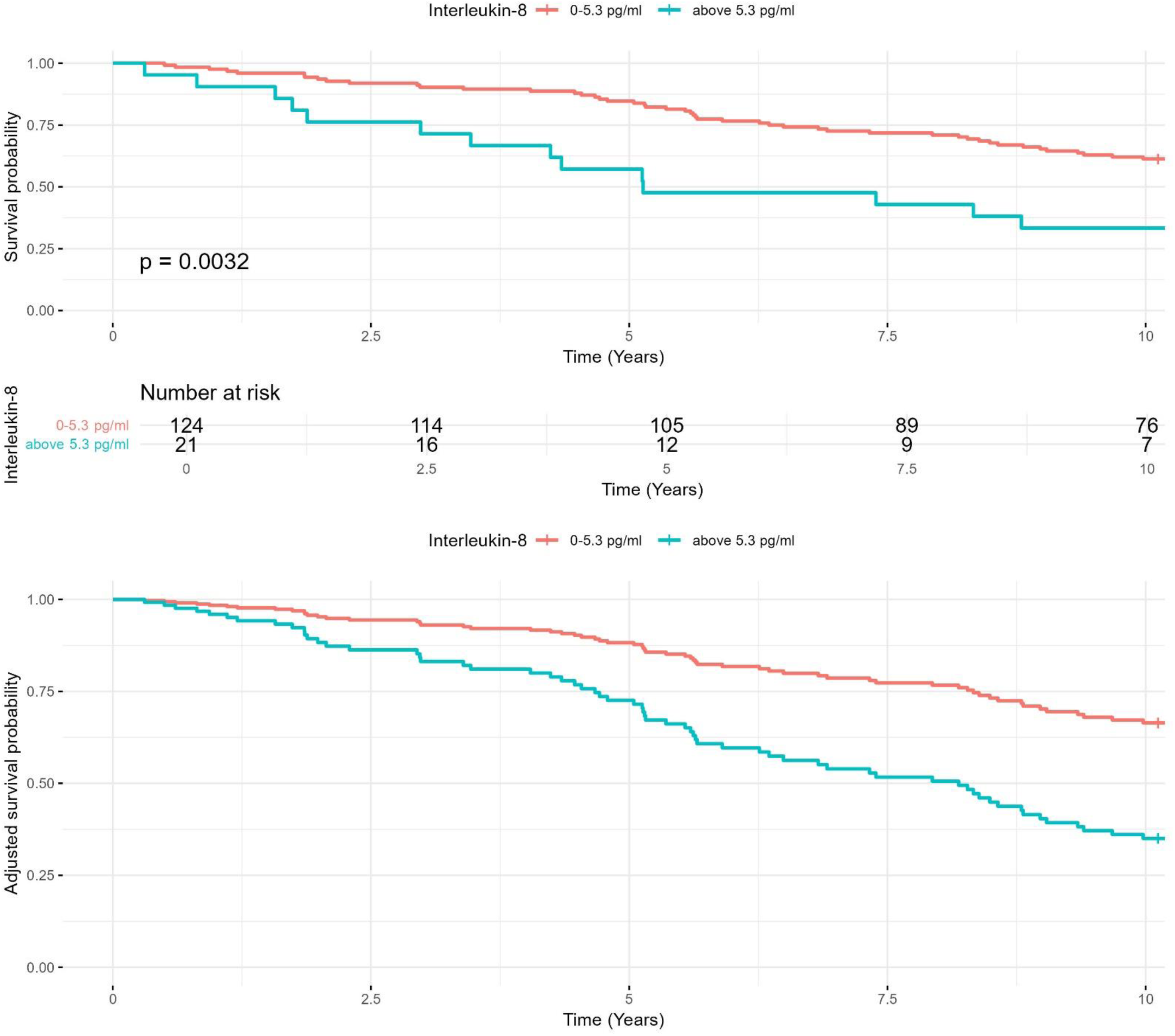
Kaplan-Meier Survival Analysis for IL-8 Levels. Kaplan-Meier survival curves comparing 10-year survival rates in heart failure patients with IL-8 levels ≤5.3 pg/ml and >5.3 pg/ml. Adjusted curves account for age, body mass index, and other clinical covariates.

### Clinical differences between IL-8 groups

Finally, we compared available clinical data of patients with IL-8 levels below detection limit (5.3 pg/ml) and above (**Table 1**). There were no significant differences regarding morphometric data, cardiovascular risk factors, pre-existing conditions, medication, or other laboratory parameters. Interestingly, patients with ↑ IL-8 levels had lower LVEF (38 % (35-45) vs. 44 % (38-48); p=0.022) and a higher percentage of patients suffering from HFrEF (62% vs. 29%; p=0.003).

## DISCUSSION

### Key findings

This post-hoc analysis of the Cognition.Matters-HF study aimed to test the hypothesis that high levels of cytokines and chemokines are independently associated with both MCI and long-term mortality in HF patients. The selection of cytokines for this study was based on their established roles in HF pathophysiology and systemic inflammation, as documented in prior research. The bead-based multiplex assay was chosen for its high sensitivity and ability to quantify multiple cytokines simultaneously, enabling a comprehensive analysis of inflammatory profiles using limited serum volumes. While the hypothesis was not supported for MCI, we observed that IL-8 levels >5.3pg/ml were independently associated with increased all-cause mortality over 10 years. These findings align with prior research suggesting a pivotal role for IL-8 in HF pathophysiology [13] and its association with adverse outcomes [14], as discussed in detail below.

### Cytokines and mild cognitive impairment

Contrary to our hypothesis and some earlier reports [4, 5], no significant association was found between cytokine levels and MCI, neither in univariable, nor in multivariable models. Our findings suggest that cytokine elevations alone may not sufficiently explain the cognitive deficits observed in HF patients. This discrepancy could be attributed to differences in study design, patient populations, or the multifactorial nature of cognitive impairment in HF, where mechanisms such as reduced cerebral perfusion [15] and neurohormonal dysregulation [16] may play a larger role. As this study was not powered to answer this hypothesis, significant associations may also have been overseen, due to inappropriate patient numbers (type II error).

We recognize that comorbid conditions, such as silent stroke, frontal disorders, and vascular dementia, may contribute to the prevalence of MCI in heart failure patients. To mitigate this, our study excluded patients with a history of clinical stroke, significant carotid artery stenosis (>50%), or psychiatric conditions such as dementia or depression. Imaging data were also reviewed when available to exclude gross structural abnormalities. However, we acknowledge that subclinical conditions, such as silent strokes or subtle vascular changes, may not have been fully accounted for and could represent alternative contributors to MCI in this cohort. This limitation underscores the multifactorial nature of MCI in HF and highlights the need for future studies with advanced imaging or biomarker-based approaches to delineate these contributions more precisely.

### Cytokines and mortality

In our study, only the elevation of IL-8 demonstrated an independent association with mortality, while other cytokines, which are commonly implicated in HF pathophysiology, including IL-6, TNF-α, and MCP-1 [2], did not. This may reflect the highly interconnected nature of the cytokine network, where overlapping and redundant pathways complicate the isolation of individual effects. Additionally, many cytokines, such as TNF-α, are more relevant in acute inflammatory responses, whereas our cohort predominantly consisted of stable chronic HF patients [17]. Moreover, the characteristics of this specific cohort, which was heterogenic with regard to etiology and phenotype of HF, may have influenced our findings. Detection limitations may have also contributed, as several cytokines had a high proportion of values below the detection threshold. Furthermore, cytokines like IL-6 and TNF-α may exert their effects indirectly or through interactions with other pathways, reducing their apparent influence in the multivariable models applied here. The observed differences in cytokine profiles likely reflect the heterogeneity of heart failure pathophysiology. Factors such as variations in systemic inflammation, cardiac remodeling, comorbid conditions, and patient-specific immune responses may all contribute to the cytokine levels. The highly interconnected and redundant nature of cytokine signaling further complicates the interpretation of these differences. While our study focused on the prognostic implications of individual cytokines, further research is needed to disentangle the complex mechanisms driving these variations.

### Interleukin-8 in heart failure

Of a variety of cytokines and chemokines are involved in HF. IL-8, also referred to as chemokine C-X-C motif ligand 8 (CXCL8), is a key chemokine primarily responsible for recruiting and activating neutrophils and monocytes [18]. As main finding of this study, we provide evidence for a prognostic impact of serum IL-8, which aligns with growing evidence supporting the role of IL-8 in HF pathophysiology [13]. Importantly, this effect was independent from clinical characteristics and HF parameters such as LVEF. While IL-6 and TNF-α have traditionally been at the forefront of inflammatory research in HF [19], IL-8 has gained attention for its specific contributions to neutrophil-driven myocardial inflammation and tissue damage [20]. We here can only speculate on the source of circulation IL-8 in HF patients. Of note, macrophages expressing IL-8 have been described in both failing and infarcted human hearts [21].

While this is the second report on the prognostic impact of IL-8 in HF [14], similar observations have been made in patients with acute myocardial infarction (AMI). In 258 AMI patients, a Norwegian study found that high IL-8 levels are associated with larger infarct sizes, reduced LVEF, adverse left ventricular remodeling, and worse clinical outcomes [22]. Another study from New Zealand included 317 AMI patients demonstrated that an IL-8 score predicted major adverse cardiovascular events [23]. Moreover, patients with acute coronary syndrome have been shown to exhibit higher IL-8 levels compared to those with stable coronary artery disease and healthy controls [24].

In our study, high levels of IL-8 associated with lower LVEF, paralleling findings from AMI patients [22]. This correlation suggests that higher IL-8 levels are indeed linked to impaired cardiac function. Pathophysiologically, IL-8 may contribute to HF progression by promoting the recruitment and activation of neutrophils and monocytes into the myocardial tissue, leading to increased inflammation, tissue damage, and adverse cardiac remodeling. This might also explain the observed association with all-cause mortality. In addition, persistent and excessive IL-8 release may also contribute to the destabilization of atherosclerotic plaques [25], potentially increasing the risk of atherothrombotic events.

Therapeutic strategies to mitigate IL-8’s effects are under clinical exploration [26]. In an AMI rat model monoclonal antibodies against IL-8 have been shown to reduce infarction size by >50% [27]. Thus, in HF, blocking IL-8 or its receptor (CXCR2) could also offer a dual benefit by reducing inflammatory damage and improving myocardial recovery. In patients with metastatic or unresectable solid tumors, the application of anti-IL-8 antibody was safe and well tolerated [28]. However, there are currently no clinical studies for cardiovascular indications and current research remains preliminary not least due to the absence of IL-8 in mice, with further investigation needed to validate anti-IL-8 approaches in clinical settings.

Interestingly, despite significant differences in IL-8 levels, C-reactive protein (CRP) levels did not vary significantly between groups. This may reflect the distinct roles of these inflammatory markers. CRP is primarily induced by interleukin-6 and represents a systemic inflammatory response, while IL-8 is more specifically involved in neutrophil recruitment and localized inflammation. The lack of correlation between these markers suggests that IL-8 may indicate specific inflammatory processes in HF, which are not captured by broader markers like CRP. This divergence highlights the complexity of HF-related inflammation and supports the utility of cytokine-specific profiling for understanding its pathophysiology.

The findings also highlight IL-8’s potential as a biomarker for long-term mortality in HF, emphasizing the role of inflammation in risk stratification. Future studies should explore IL-8’s utility as a therapeutic target and its integration into multimodal approaches combining biomarkers, imaging, and genetic analyses to refine HF management.

### Strenghts and limitations

Our study’s strengths include the use of a well-characterized HF cohort with long-term follow-up and robust multivariable modeling to account for confounding factors.

A key limitation of this study is its cross-sectional design, which captures cytokine levels at a single time point. Cytokines and HF both exhibit significant temporal variability, influenced by disease progression, treatment changes, and other factors. Consequently, the observed associations between cytokine concentrations and long-term outcomes may not fully reflect their dynamic relationship. Longitudinal studies evaluating time-curve changes in cytokine levels are essential to better understand their prognostic value and potential as therapeutic targets in HF.

Limitations also include the small sample size, which may increase the risk of Type II errors. Differential blood counts were not available. Additionally, the high proportion of cytokine values below detection limits in the multiplex assay could have reduced sensitivity to detect associations with less abundant markers. This also required dichotomization of cytokine levels using manufacturer-provided detection limits as cut-offs, which may introduce bias and reduce the granularity of the findings. Future studies with larger sample sizes and improved sensitivity of cytokine assays are needed to validate these findings and explore alternative analytical approaches, such as deriving cut-offs using receiver operating characteristic curve analysis.

Further, a limitation of this study is that the HF cohort did not receive sodium-glucose cotransporter 2 inhibitors, glucagon-like peptide-1 receptor agonists, or angiotensin receptor-neprilysin inhibitors, as recruitment occurred between 2011 and 2014, prior to their clinical adoption. While reflecting standard practice at the time, this limits the applicability of our findings to current HF management. Future studies should evaluate how these therapies might influence inflammatory profiles and outcomes.

An additional limitation of this study is that the exclusion criteria did not account for inflammatory comorbidities, recent infections, or trauma, which could have influenced cytokine levels. Although all participants had clinically stable HF at the time of recruitment, these factors may contribute to variability in inflammatory markers and should be addressed in future studies.

Further, although our study did not find an association between cytokine profiles and MCI, it is plausible that inflammation, rather than HF itself, may underlie MCI in these patients. The lack of association might reflect the multifactorial etiology of MCI, where systemic inflammation could play a contributing role independent of HF.

### Conclusion

This post-hoc cytokine analysis reinforces the pivotal role of inflammation in HF progression, with high IL-8 levels independently predicting long-term mortality. These findings highlight IL-8’s potential as a prognostic biomarker and therapeutic target. The absence of associations with other cytokines underscores the complexity of HF’s inflammatory pathways, while IL-8’s specific role suggests a unique contribution to adverse outcomes. Further research is warranted to validate these results and assess targeted interventions against IL-8 in HF.

## Data Availability

All data produced in the present study are available upon reasonable request to the authors

## INSTITUTIONAL REVIEW BOARD STATEMENT

The study was conducted in accordance with the Declaration of Helsinki, and approved by the Ethics Committee of the University of Würzburg (protocol code #245/10) on 17 January 2011.

## INFORMED CONSENT STATEMENT

Informed consent was obtained from all subjects involved in the study.

## DATA AVAILABILITY STATEMENT

The data presented in this study are available on request from the corresponding author.

## ACKNOWLEDGMENTS

We would like to thank the interdisciplinary Cognition.Matters-HF team for the excellent conduction of the study and the lab of Michael Schuhmann for their excellent technical support.

